# Assessing the Impact of Downsampled ECGs and Alternative Loss Functions in Multi-Label Classification of 12-Lead ECGs

**DOI:** 10.1101/2022.11.16.22282373

**Authors:** Bjørn-Jostein Singstad, Eraraya Morenzo Muten

## Abstract

The electrocardiogram (ECG) is an almost universally accessible diagnostic tool for heart disease. An ECG is measured by using an electrocardiograph, and today’s electrocardiographs use built-in software to interpret the ECGs automatically after they are recorded. However, these algorithms show limited performance, and therefore clinicians usually have to manually interpret the ECG, regardless of whether an algorithm has interpreted the ECG or not. Manual interpretation of the ECG can be time-consuming and require specific skills. Therefore, a better algorithm is clearly needed to make correct ECG interpretations more accessible and time efficient. Algorithms based on artificial intelligence have shown promising performance in many fields, including ECG interpretation, over the last few years and might represent an alternative to manual ECG interpretation.

In this study, we used a dataset with 88253 12-lead ECGs from multiple databases, annotated with SNOMED-CT codes by medical experts. We employed a supervised convolutional neural network with an Inception architecture to classify 30 of the most frequent annotated diagnoses in the dataset. Each patient could have more than one diagnosis, which makes this a multi-label classification. We compared the Inception model’s performance while applying different preprocessing methods on the ECGs and different model settings during 10-folded cross-validation. We compared the model’s classification performance using binary cross-entropy (BCE) loss and double soft F1 loss. Furthermore, we compared the classification performance when downsampling the original sampling rate of the input ECG. Finally, we trained 30 interpretable linear models to provide class activation maps to explain the relative importance of each sample in the ECG with respect to the 30 diagnoses considered in this study.

Due to the heavily imbalanced class distribution in our dataset, we placed the most emphasis on the F1 score when evaluating the performance of the models. Our results show that the best performance in terms of F1-score was seen when the Inception model used double soft F1 as the loss function and ECGs downsampled to 75Hz. This model achieved an F1 score of 0.420 ± 0.017, accuracy = 0.954 ± 0.002, and an AUROC score of 0.832 ± 0.019. An aggregation of the generated saliency maps, achieved using Local Interpretable Model-Agnostic Explanations (LIME), showed that the Inception model paid the most attention to the limb leads and the augmented leads and less importance to the precordial leads.

One of the more significant contributions that emerge from this study is the use of aggregated saliency maps to obtain ECG lead importance for different diagnoses. In addition, we emphasized the relevance of evaluating different loss functions, and in this specific case, we found double soft F1 loss to be slightly better than BCE. Finally, we found it somewhat surprising that downsampling the ECG led to higher performance compared to the original 500Hz sampling rate. These findings contribute in several ways to our understanding of the artificial intelligence-based interpretation of ECGs, but further studies should be carried out to validate these findings in other datasets from other patient cohorts.

## 1 Introduction

Cardiovascular disease (CVD) is one of the leading causes of death worldwide. Numbers from World Health Organization estimate that 17.9 million people died from CVD in 2016 which represented 31% of all global deaths that year [1]. Early detection of patients with a risk of CVD could potentially reduce the severity of the disease and also decrease the number of persons who die from CVD.

Electrocardiography is a non-invasive and widely used method to record electrocardiograms (ECG), which has enabled clinicians to interpret, diagnose and prognosticate heart disease since the beginning of the 20th century [2]. The ECG is the result of a measurement of the electrical activity of the heart by recording the voltage potential from electrodes placed on the patient’s skin. Electrocardiography is generally easier to set up and more cost-effective compared to other diagnostic methods such as echocardiogram and magnetic resonance imaging of the heart. On the other hand, one of the challenges is that the ECG can be difficult to interpret correctly. Correct interpretation can be time-consuming and require a high degree of expertise [3].

In the 1950s it became possible to convert analog ECG signals to digital format and this led to the development of digital interpretation algorithms in the 1960s [4]. Today, most of the modern and clinically used electrocardiographs are equipped with built-in interpretation software. The software interprets the ECG and prints interpretive texts that may indicate different pathologies. Studies show that there are several limitations to the automatic interpretation algorithms [5, 4]. The errors, done by the automatic interpretation algorithms, imply that doctors have to read over the ECGs to ensure that the diagnosis is correct. This is time-consuming for the doctors and leads to high interpretation variability. Thus, there is a need for developing a better ECG interpretation algorithm, since this may lead to less time-consuming interpretation for the doctors, less variability in the interpretation and better diagnostic performance which may lead to earlier detection and treatment of patients with CVD.

In the past decades, several new important trends have converged and may potentially be ushering in a new age with significance to ECG interpretation. Firstly, ECGs are now increasingly stored in digital format, allowing computerized analysis of massive data sets. Secondly, personal sensors such as training monitors and smartwatches (e.g., Apple Watch, Withings Watch, Samsung Galaxy Watch) now include simple ECG recording abilities, further expanding access to ECGs and the range of people studied. Finally, artificial intelligence (AI) or more specifically deep learning (DL) has shown remarkable abilities in classifying signal data [6], and more specifically also ECG data [7, 8, 9, 10, 11, 12, 13].

Despite the good performance of the DL-based ECG interpretation models, the doctors are still responsible for the diagnosis, and such models should then be considered as decision support tools, but the complexity in DL models makes the decision inaccessible to humans, often referred to as the black box phenomenon [14]. This has led to the development of another sub-field within AI, explainable AI (XAI) [15], with the aim of making the model decision more human-interpretable. XAI methods such as Gradient class activation map (GradCAM) [16], LIME [17] and SHAP [18] have already been used to get class activation maps, showing which part of the raw ECG waveform was most important for the DL model’s prediction. The majority of these studies have focused on explaining single-label classification models [19, 20, 21, 22], while only a few have explained multi-label classification models [23, 24]. In one study, the researchers discovered novel disease-specific ECG features in Phospholamban (PLN) mutation carriers [19], but for many other diseases, the DL model will likely rely on very subtle patterns and combinations of features from different leads, and even though these get highlighted and displayed to the doctors in the class activation map, it might be hard for them to understand or verify the relationship between the features used by the DL-model.

This study builds on the George Moody Challenge 2020 [25] and 2021 [26] where the objective was to perform multi-label classification of cardiovascular diagnoses using the raw ECG waveform. We contributed to the 2020 edition of the George Moody challenge by combining convolutional neural networks and rule-based algorithms [27] and in the 2021 edition, we used classifier chains based on convolutional neural networks [28]. In the current study, we compare inception models trained using BCE and double soft F1-loss and show how the sampling frequency of the ECG records affects the classification performance. Furthermore, we use explainable AI techniques to investigate which of the 12 leads has the highest importance when classifying different diagnoses.

## 2 Methods and materials

### 2.1 Data

We used ECG data from seven different open-access databases [26, 25, 29, 30, 31, 32, 33], with a total of 88253 12-lead ECGs in waveform format. All ECGs different from 10 seconds in recording length were excluded, and 81327 ECGs were used for further development and validation as Figure 1 shows. Each ECG was stored in a .mat file and had a corresponding .hea file containing metadata such as the ECG recording length, sample frequency, the patient’s age, gender and diagnosis. There was a total of 133 different experts annotated diagnoses in the dataset, but in this study, we choose to consider only 30 of them (the same 30 used in George Moody PhysioNet Challenge 2021 [26]). The prevalence of each of these 30 diagnoses are shown in Table 1. Each patient could have more than one of the 30 diagnoses at the same time, which makes this task a multi-label classification task with more than 3000 different combinations of diagnoses among the patients in the dataset.

**Figure 1:**
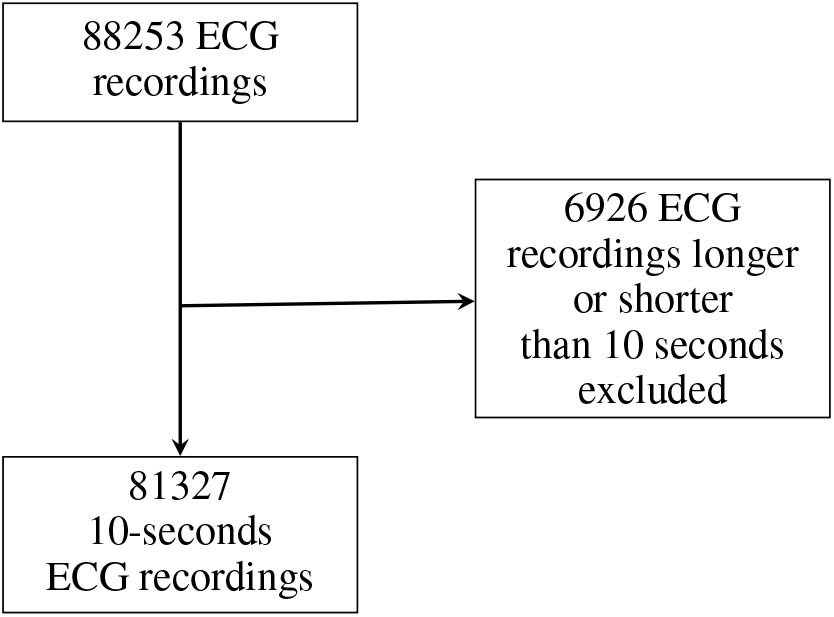
Patients with an ECG recording shorter or longer than 10 seconds were excluded from the development set. 6926 ECGs were excluded and the remaining 81327 ECGs were used to train and validate the model.

**Table 1:** The prevalence of the 30 classified diagnoses after excluding patients with an ECG recording different from 10 seconds.

| Diagnoses | Prevalence | Diagnoses | Prevalence |
| --- | --- | --- | --- |
| 1st Degree AV block | 3291 | Premature Atrial Contraction | 2827 |
| Atrial Fibrillation | 5062 | Premature Ventricular Contractions | 1259 |
| Atrial Flutter | 8509 | Prolonged PR Interval | 393 |
| Bradycardia | 267 | Prolonged QT Interval | 2152 |
| Bundle Branch Block | 511 | Q Wave Abnormal | 2261 |
| Complete Left Bundle Branch Block | 218 | Right Axis Deviation | 1482 |
| Complete Right Bundle Branch Block | 2015 | Right Bundle Branch Block | 2331 |
| Incomplete Right Bundle Branch Block | 2306 | Sinus Arrhythmia | 4176 |
| Left Anterior Fascicular Block | 2665 | Sinus Bradycardia | 19303 |
| Left Axis Deviation | 7952 | Sinus Rhythm | 30426 |
| Left Bundle Branch Block | 1260 | Sinus Tachycardia | 10261 |
| Low QRS Voltage | 1765 | Supraventricular Premature Beats | 267 |
| Nonspecific Intraventricular Conduction Disorder | 2101 | T Wave Abnormal | 12673 |
| Pacing Rhythm | 1694 | T Wave Inversion | 4340 |
| Poor R Wave Progression | 656 | Ventricular Premature Beats | 731 |

### 2.2 Preprocessing

#### 2.2.1 ECG processing

More than 85% of all ECGs in the development set were initially sampled at 500 Hz. All ECGs were resampled to the same sample frequency. In this study, we compared the model’s performance when the signal was downsampled from 500 to 400, 300, 200, 100, 75, 50 and 25Hz.

#### 2.2.2 Label processing

We one-hot encoded all the 30 diagnoses considered in this study, such that each ECG recording had a corresponding 30-bit long array of ones or zeros. A binary one means that the patient has the given diagnosis and zero means that the patient does not have the diagnosis.

### 2.3 CNN architecture

We developed a CNN model inspired by the Inception architecture [34] as shown in Figure 2 using TensorFlow [35]. The input to this model was an array, representing the raw ECG. The array containing the ECG signal can be denoted as:

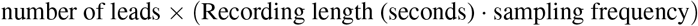

**Figure 2:**
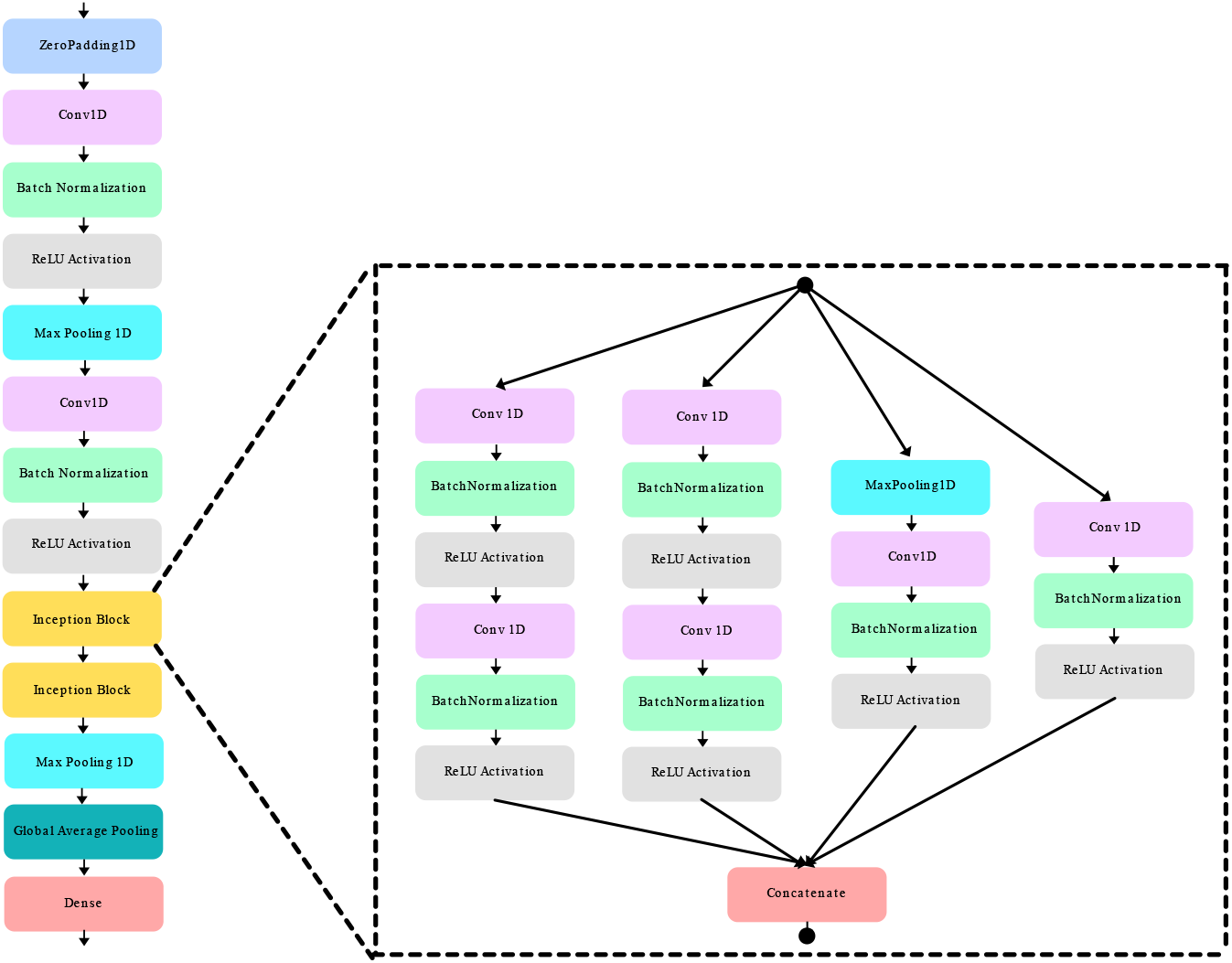
Inception model architecture. Each block represents a mathematical operation in the convolutional neural network. The blocks inside the dashed lines represent an Inception block.

The output layer of the model had 30 neurons, corresponding to the 30 scored diagnoses. A Sigmoid activation was used in the final layer, giving a continuous number between 0 and 1 for each of the 30 diagnoses.

### 2.4 Loss function

A loss function is used to compute the error of the prediction made by the model during the training phase. The computed errors are used to adjust the weight coefficient in the model using backpropagation [36]. A previous study claimed that different variations of F soft loss could be beneficial when performing multi-label classification with imbalanced classes [37]. In this study, we compared the Inception model using double soft F1 loss and binary cross-entropy loss.

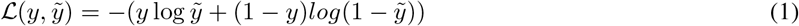

Equation 2 shows how double soft F1-loss 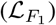 is calculated. The small number (+10^*−*16^) is added to the denominator to prevent the function to divide by zero.

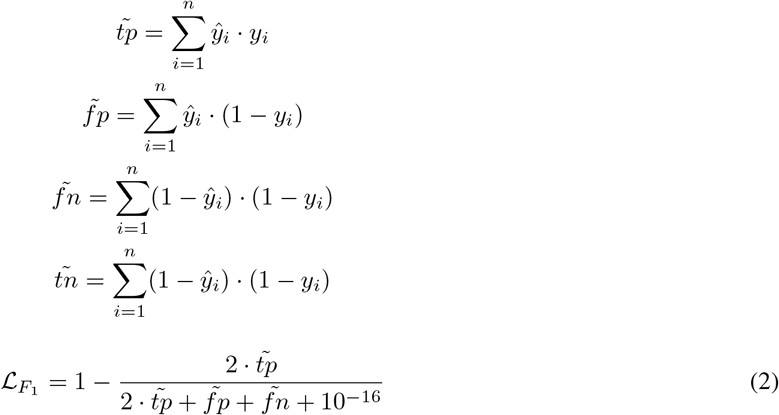

An Adam optimizer was used to compute the gradients, based on the loss, which was backpropagated to update the weights of the artificial neurons in the Inception model [38].

### 2.5 Training and validation

The model was trained and evaluated on the dataset using 10-fold stratified cross-validation. The data were stratified based on the prevalence of the diagnoses to ensure similar distribution of diagnoses in both the train and validation fold. The models were trained for 15 epochs, with a batch size of 30 and a learning rate of 0.001.

The model performance was scored using the area under the curve (AUC) of the receiver operating characteristic (ROC) curve, F1-score and average accuracy across all classes (hereby just referred to as accuracy). Equation 3 shows how we compute accuracy by comparing the true label (*y*) and the predicted label (*ŷ*) for each ECG recording, *n*_*s*_ and then finding the average accuracy for each class *c* and finally taking the average across all classes, *n*_*c*_.

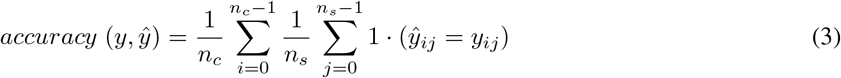

### 2.6 Explainability

To find the relative importance of the features in the ECGs, a local interpretive model-agnostic explanation (LIME) model was trained to fit the input data (ECGs) to the output predictions from the Inception model. A LIME model is a linear surrogate model which is easier to interpret compared to a deep neural network. One LIME model was trained and tested for each of the 30 classes. As output, a LIME model provides a class activation map that has an equal shape to the input. Values close to zero in the class activation map mean low activation, while higher values mean higher activation. Figure 3 shows an example of one ECG-lead (aVL), visualized in the same plot as the corresponding activation map for that specific lead. The LIME model is here trained to explain the atrial fibrillation classification from the Inception model. The dark red lines indicate high activation, while the brighter color indicates lower activation. This example is what’s called a local explanation because it only explains the model’s behavior on a single input data, whereas global explanations are used to explain the model’s behavior on the whole population.

**Figure 3:**
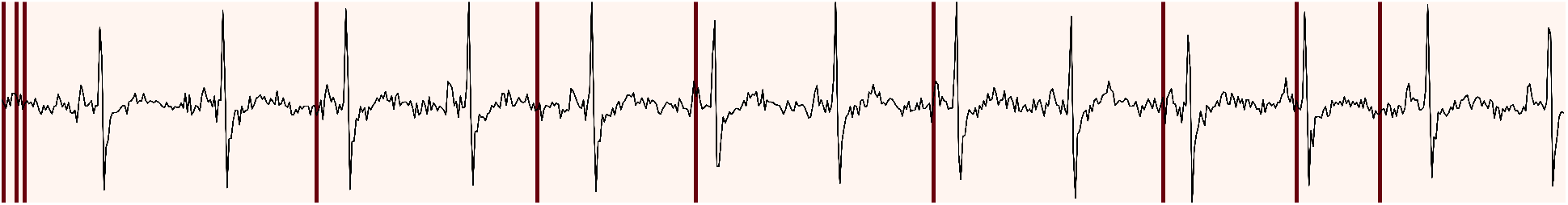
Lead aVL visualized together with its class activation map for a patient correctly classified with atrial fibrillation with a probability of 0.998. The red vertical lines show the features in the ECG that contributed most towards the prediction, according to the local interpretable model-agnostic explanation (LIME) model.

#### 2.6.1 Developing the LIME model

We trained a LIME model for each of the 30 classes using the training data from the 10th cross-validation split during the model development. For each LIME model, 1000 ECGs labeled with the class to explain and 1000 ECGs labeled with classes different from the one to explain were used to train the LIME model. The trained LIME models were then applied on the ECGs in the validation split of the 10th cross-validation split. The *n*-th LIME model was applied on all ECGs labeled with the *n*-th class in the validation split.

## 3 Results

### 3.1 Loss function

The box plots in Figure 4 compare the performance of the Inception model trained using BCE loss with the Inception Time model trained using double soft F1-loss. Each box represents the ten values achieved during the 10-folded cross-validation. Figure 4a shows the achieved accuracy, Figure 4b shows the F1-score and Figure 4c shows the AUROC score.

**Figure 4:**
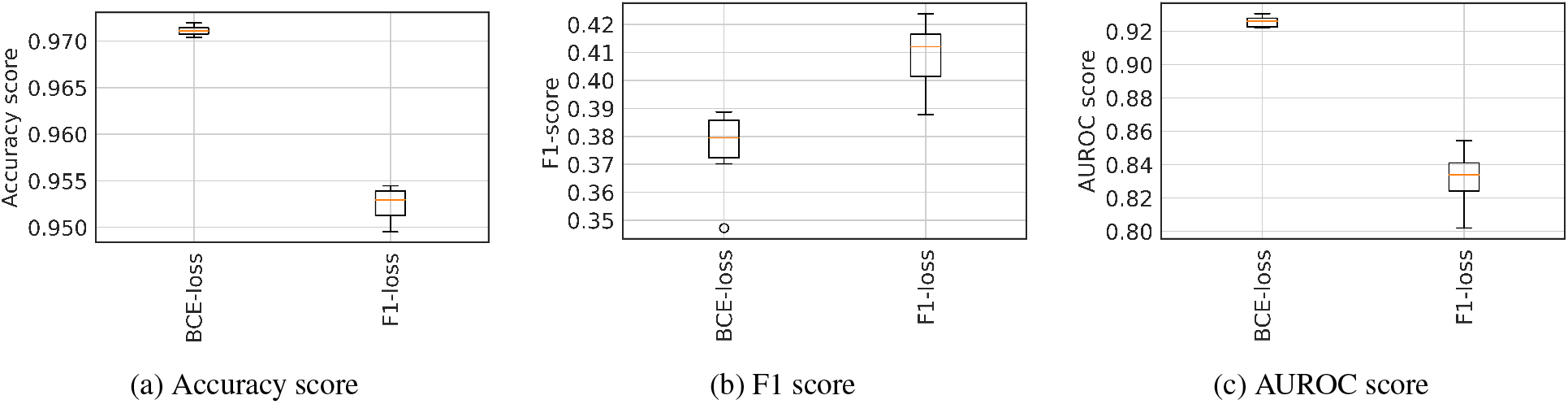
Classification performance achieved by the Inception model trained on 12-lead ECGs, using binary cross-entropy loss and double soft F1-loss. The box plots are accumulated scores obtained on the validation folds during 10-folded cross-validation. (a) shows the accuracy scores, (b) shows the F1-scores and (c) shows the AUROC scores.

Due to the heavily imbalanced dataset used in this study, we selected the loss function that achieved the best F1-score, which was double soft F1-loss.

### 3.2 Sampling frequency

In order to assess the impact of the ECGs sampling frequency on the classification results, we took eight copies of the original dataset and resampled the datasets to eight different sample frequencies (25Hz, 50Hz, 75Hz, 100Hz, 200Hz, 300Hz, 400Hz, 500Hz). Eight Inception models, using double soft F1-loss, were trained and validated using 10-fold CV. All eight models were trained for 15 epochs, with a batch size of 30, using Adam as the optimizer and an initial learning rate of 0.001. In Figure 5 we compare the cross-validated accuracy, F1-score and AUROC score obtained by the eight models.

**Figure 5:**
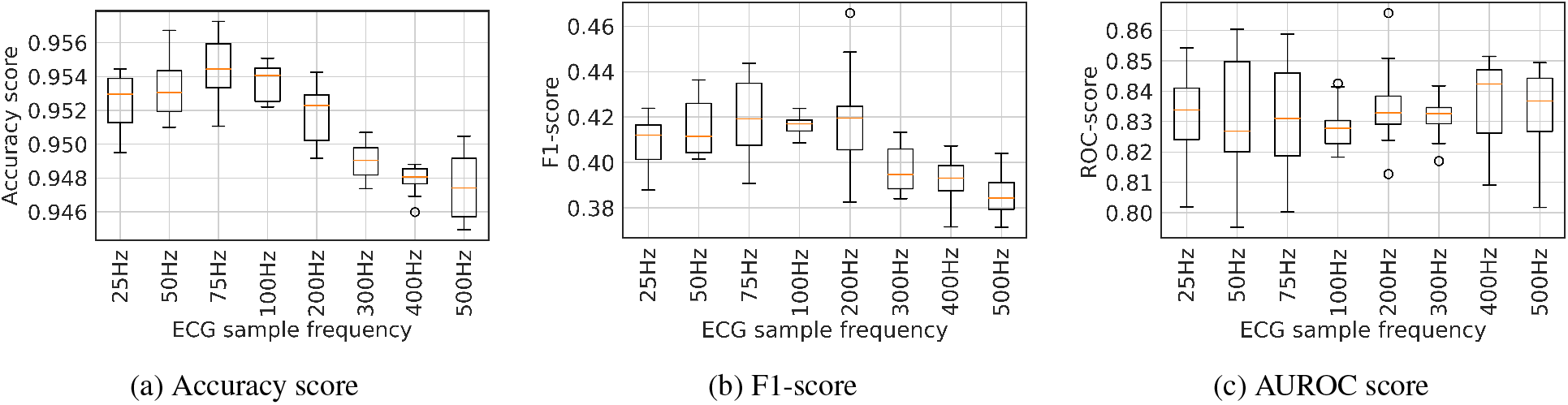
Classification results performed on the ECGs at different sampling frequencies. Each box represents the accumulated scores obtained on the validation folds by the Inception model, using double soft F1-loss, during 10-folded cross-validation. (a) shows the accuracy score, (b) shows the F1-score and (c) shows the AUROC scores.

### 3.3 Explainability

Finally, an Inception model, with a double soft F1-score as the loss function was trained on ECG signals resampled to 75Hz. The Inception model was trained on the training data from the 10th split of the 10-folded CV and tested on the validation fold. 30 LIME models were trained and tested for each of the 30 diagnoses. Figure 6 shows a saliency map of the ECG leads with the highest activation/importance for each of the 30 diagnoses. The saliency map was obtained by finding the ECG lead with the highest average activation value for each of the 30 diagnoses in the 10th validation fold.

**Figure 6:**
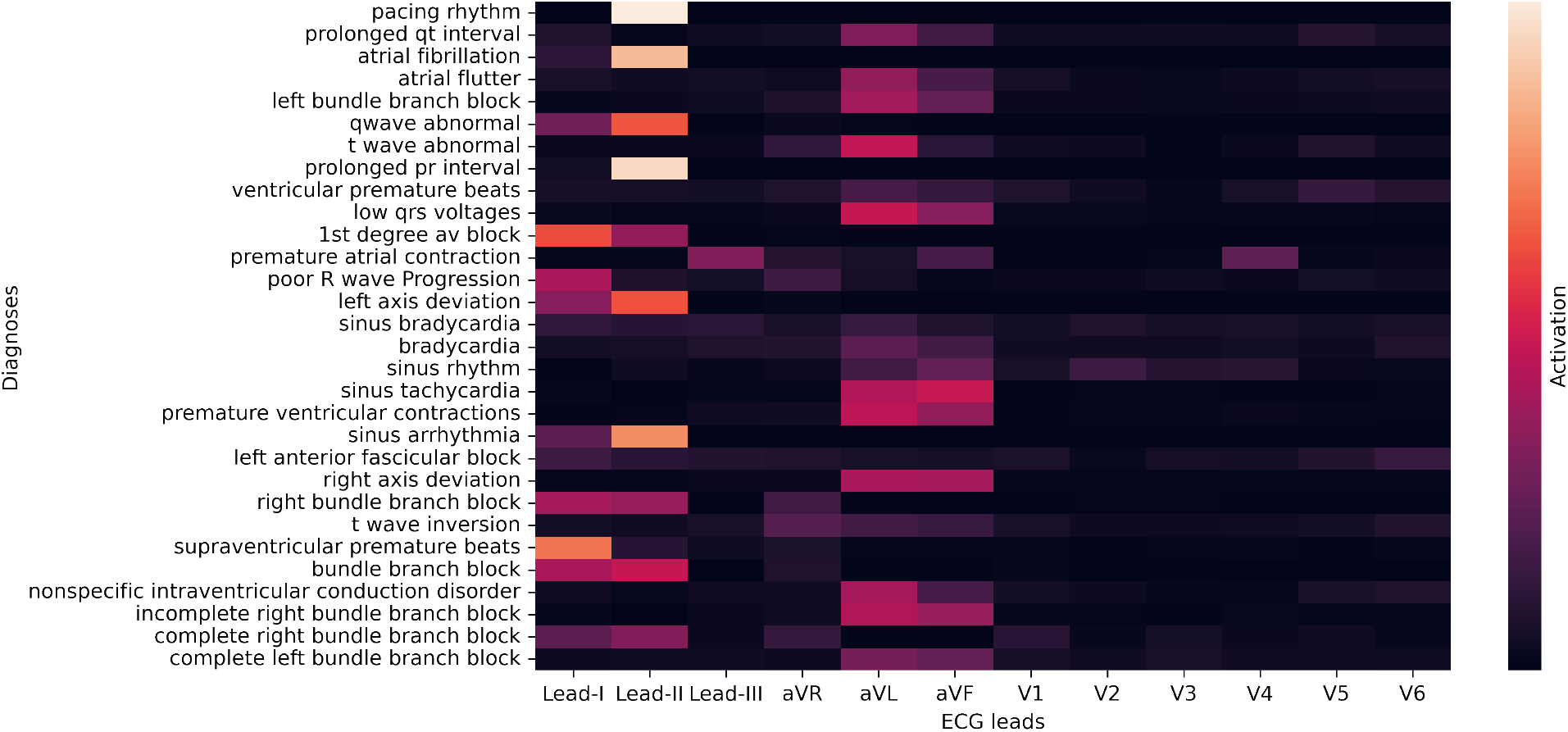
Aggregated results from the class activation maps provided by the LIME model when interpreting the predictions from the Inception model that is interpreting the validation data. The x-axis shows the 12 ECG leads and the y-axis represents the 30 different diagnoses represented in the dataset. Dark colors in the saliency map mean low activation, while bright means high activation.

## 4 Discussion

This study demonstrates an Inception-type convolutional neural network doing multi-label classification on an imbalanced ECG dataset. Additionally, we also employed an explainable AI technique called LIME in order to find the ECG lead with the highest class activation for each of the 30 diagnoses considered in this study. To the best of our knowledge, this is the first time class activation maps have been used to determine ECG lead importance for different diagnoses.

During development of the Inception model, we compared two different loss functions, BCE and double soft F1-loss. We found that double soft F1-loss gave a significantly better F1-score (Figure 4b), which is considered the most important metric in a heavily imbalanced dataset such as the one used in this study. It is however somewhat surprising that the model using BCE loss achieved better accuracy and AUROC score than the model using double soft F1-loss. A plausible explanation seems to be that the BCE model was good at classifying the major classes, giving a high accuracy score, and the model using double soft F1-loss was generally good at classifying all 30 classes, giving a high F1-score.

One of the most surprising findings in this study was the improvement in classification performance when downsampling the original 500 Hz ECGs. As seen in Figure 5 both the accuracy (Figure 5a) and F1-score (Figure 5b) performance reach their peak around 75Hz. The increase in classification performance using downsampled ECG signals could be a bit counter-intuitive since one would expect the ECG to lose a lot of important information. A possible explanation for this is that there is an ideal ratio between convolution kernel size and the features in the ECG, such as P-waves T-waves and QRS complex. However, we also did some experiments by increasing the kernel size, but this did not give the same improvement in classification performance as lowering the ECG sampling frequency. Nonetheless, this needs more research to reach a concusion.

The saliency map in Figure 6 is an aggregation of all class activation maps achieved from the 10th cross-validation fold. The figure shows that lead II is the lead with the highest overall activation across all diagnoses, and the precordial leads (V1-V6) generally show low activation. A possible explanation for this is that lead II often has a high signal-to-noise ratio compared to the other leads. In addition, the majority of diagnoses considered in this study are arrhythmias and these diagnoses are generally diagnosed by looking at the limb leads by human interpreters also.

### 4.1 Augmentation

Augmentation has shown promising effects in various image classification tasks [39]. Therefore we hypothesized that augmentation might have a good effect on signal and ECG classification as well. More specifically, we tried to add random noise and baseline wander to the ECG signals. However, these augmentations did not significantly improve the performance of our models.

The random noise was induced by adding a random number (*N*) in the range of the ± standard deviation (*σ*) of all values in the current ECG recording, shown in Equation 4.

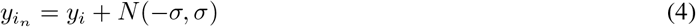

Baseline wander was induced to the signal by adding a cosine wave from 0 to 2*π* and shifting the cosine wave randomly between 0 and 2*π*. The amplitude of the signal was randomly set by multiplying a random number (N) drawn from the distribution of all values in an ECG recording, shown in Equation 5.

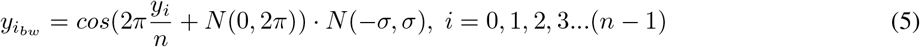

Figure 7a shows an example of an unprocessed ECG and Figure 7b shows the same ECG with added random noise using the method described in Equation 4. Figure 7c shows the ECG in Figure 7a with simulated baseline wander as described in Equation 5.

**Figure 7:**
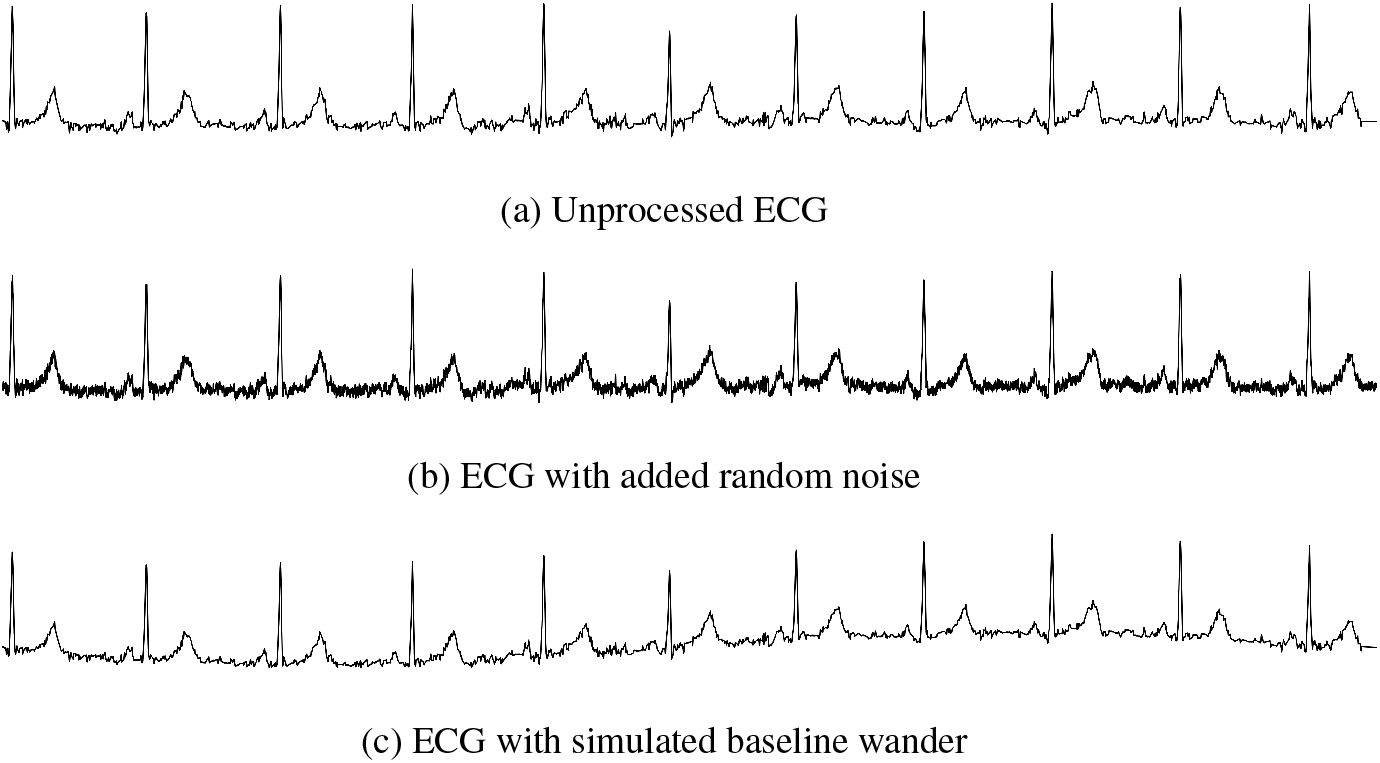
Comparing an unprocessed ECG (a) with the same ECG with added noise (b) and simulated baseline wander (c)

### 4.2 Limitation

One key limitation of the results in this work is that we did not test the model on a separate and independent test set. The selection of optimal loss function and sampling rate could therefore have resulted in overfitting to the present dataset. However, the datasets used to train and validate the model are from different hospitals across the world and is therefore likely to represent a great diversity of patients, but anyway, validation on an external test set is needed to control the model for potential overfitting that could have occurred in our study.

In order to create the class activation maps and the aggregated lead importance diagram (Figure 6) we used the LIME framework. One of the limitations of this approach is that the LIME module we used was originally intended to be used on recurrent neural networks and not convolutional neural networks as done in this study. Secondly, previous research has stated that methods such as LIME are too generic and should be used with care on waveform data [40]. Comparing activation maps from LIME with model-specific explanation methods, such as Grad-CAM [16], would therefore be interesting

### 4.3 Future perspectives

Future studies should consider other loss functions than binary cross-entropy when training neural network-based multi-label classification models on imbalanced datasets. Also, one should assess different ECG sampling frequencies to get optimal performance. In terms of model explainability, future studies should let medical doctors or cardiologists verify the ECG activation maps to assess the usefulness of the XAI.

## 5 Conclusion

The primary aim of this study was to train a multi-label ECG classifier to achieve the best possible performance, given the unbalanced dataset score. Furthermore, we used this model to obtain class activation maps and based on those we found the leads that were considered the most important for each diagnosis. We also found that double soft F1-loss might improve the performance when classifying heavy imbalanced datasets. In addition, we observed that reducing the sampling frequency of the ECG from 500 Hz to around 75 Hz increased the model performance.

## Data Availability

All data produced are available online at: https://github.com/Bsingstad/Post-George-Moody-challenge-2020-2021

## Code availability

The complete source code described in this paper is openly available on GitHub (https://github.com/Bsingstad/Post-George-Moody-challenge-2020-2021) under a free software license (CC-BY 4.0).

## Acknowledgment

We would like to thank Antony M. Gitau for proofreading and giving constructive criticism of the manuscript.

## References

[1] WHO Cardiovascular Diseases (CVDs). en. Accessed 21.08.2022. URL: https://www.who.int/news-room/fact-sheets/detail/cardiovascular-diseases-(cvds) (visited on 08/21/2022).

[2] S. Serge Barold. “Willem Einthoven and the birth of clinical electrocardiography a hundred years ago”. eng. In: Cardiac Electrophysiology Review 7.1 (Jan. 2003), pp. 99–104. ISSN: 1385-2264. DOI: 10.1023/a:1023667812925.

[3] Martin Bickerton and Alison Pooler. “Misplaced ECG Electrodes and the Need for Continuing Training”. In: British Journal of Cardiac Nursing 14.3 (Mar. 2019), pp. 123–132. DOI: 10.12968/bjca.2019.14.3.123.

[4] Harold Smulyan. “The Computerized ECG: Friend and Foe”. In: The American Journal of Medicine 132.2 (Feb. 2019), pp. 153–160. DOI: 10.1016/j.amjmed.2018.08.025.

[5] Jürg Schläpfer and Hein J. Wellens. “Computer-Interpreted Electrocardiograms”. In: Journal of the American College of Cardiology 70.9 (Aug. 2017), pp. 1183–1192. DOI: 10.1016/j.jacc.2017.07.723.

[6] Hassan Ismail Fawaz and et al. “Deep Learning for Time Series Classification: A Review”. en. In: Data Mining and Knowledge Discovery 33.4 (July 2019),pp. 917–963. DOI: 10.1007/s10618-019-00619-1.

[7] Antônio H. Ribeiro et al. “Automatic diagnosis of the 12-lead ECG using a deep neural network”. en. In: Nature Communications 11.1 (Apr. 2020). Number: 1 Publisher: Nature Publishing Group, p. 1760. ISSN: 2041-1723. DOI: 10.1038/s41467-020-15432-4. URL: https://www.nature.com/articles/s41467-020-15432-4 (visited on 07/24/2020).

[8] Jagadeeswara Rao Annam et al. “Classification of ECG Heartbeat Arrhythmia: A Review”. en. In: Procedia Computer Science. Third International Conference on Computing and Network Communications (CoCoNet’19) 171 (Jan. 2020), pp. 679–688. ISSN: 1877-0509. DOI: 10.1016/j.procs.2020.04.074. URL: http://www.sciencedirect.com/science/article/pii/S1877050920310425 (visited on 12/16/2020).

[9] Qihang Yao et al. “Multi-class Arrhythmia detection from 12-lead varied-length ECG using Attention-based Time-Incremental Convolutional Neural Network”. en. In: Information Fusion 53 (Jan. 2020), pp. 174–182. ISSN: 15662535. DOI: 10.1016/j.inffus.2019.06.024. URL: https://linkinghub.elsevier.com/retrieve/pii/S1566253518307632 (visited on 03/03/2020).

[10] Dengao Li et al. “Automatic Classification System of Arrhythmias Using 12-Lead ECGs with a Deep Neural Network Based on an Attention Mechanism”. en. In: Symmetry 12.11 (Nov. 2020).Number: 11 Publisher: Multidisciplinary Digital Publishing Institute, p. 1827. DOI: 10.3390/sym12111827. URL: https://www.mdpi.com/2073-8994/12/11/1827 (visited on 12/16/2020).

[11] Tsai-Min Chen et al. “Detection and Classification of Cardiac Arrhythmias by a Challenge-Best Deep Learning Neural Network Model”. en. In: iScience 23.3 (Mar. 2020), p. 100886. ISSN: 25890042. DOI: 10.1016/j.isci.2020.100886. URL: https://linkinghub.elsevier.com/retrieve/pii/S2589004220300705 (visited on 03/03/2020).

[12] Zachi I Attia et al. “Application of artificial intelligence to the electrocardiogram”. In: European Heart Journal 42.46 (Dec. 2021), pp. 4717–4730. ISSN: 0195-668X. DOI: 10.1093/eurheartj/ehab649. URL: https://doi.org/10.1093/eurheartj/ehab649 (visited on 12/13/2021).

[13] Awni Y. Hannun et al. “Cardiologist-level arrhythmia detection and classification in ambulatory electrocardiograms using a deep neural network”. en. In: Nature Medicine 25.1 (Jan. 2019), pp. 65–69. ISSN: 1078-8956, 1546-170X. DOI: 10.1038/s41591-018-0268-3. URL: http://www.nature.com/articles/s41591-018-0268-3 (visited on 08/16/2020).

[14] Davide Castelvecchi. “Can we open the black box of AI?” en. In: Nature News 538.7623 (Oct. 2016). Section: News Feature, p. 20. DOI: 10.1038/538020a. URL: http://www.nature.com/news/can-we-open-the-black-box-of-ai-1.20731 (visited on 12/18/2020).

[15] Arun Rai. “Explainable AI: from black box to glass box”. en. In: Journal of the Academy of Marketing Science 48.1 (Jan. 2020), pp. 137–141. ISSN: 1552-7824. DOI: 10.1007/s11747-019-00710-5. URL: https://doi.org/10.1007/s11747-019-00710-5 (visited on 12/04/2021).

[16] Ramprasaath R. Selvaraju et al. “Grad-CAM: Visual Explanations from Deep Networks via Gradient-Based Localization”. In: 2017 IEEE International Conference on Computer Vision (ICCV). ISSN: 2380-7504. Oct. 2017, pp. 618–626. DOI: 10.1109/ICCV.2017.74.

[17] Marco Tulio Ribeiro, Sameer Singh, and Carlos Guestrin. ““Why Should I Trust You?”: Explaining the Predictions of Any Classifier”. In: 1602.04938 [cs, stat] (Aug. 2016). arXiv: 1602.04938. URL: http://arxiv.org/abs/1602.04938 (visited on 08/27/2020).

[18] Scott M Lundberg and Su-In Lee. “A Unified Approach to Interpreting Model Predictions”. In: Advances in Neural Information Processing Systems 30. Ed. by I. Guyon et al. Curran Associates, Inc., 2017, pp. 4765–4774. URL: http://papers.nips.cc/paper/7062-a-unified-approach-to-interpreting-model-predictions.pdf.

[19] van de Leur Rutger R. et al. “Automatic Triage of 12-Lead ECGs Using Deep Convolutional Neural Networks”. In: Journal of the American Heart Association 9.10 (May 2020). Publisher: American Heart Association, e015138. DOI: 10.1161/JAHA.119.015138. URL: https://www.ahajournals.org/doi/10.1161/JAHA.119. 015138 (visited on 01/09/2021).

[20] Steven A. Hicks and et al. “Explaining deep neural networks for knowledge discovery in electrocardiogram analysis”. en. In: Scientific Reports 11.1 (May 2021). Bandiera_abtest: a Cc_license_type: cc_by Cg_type: Nature Research Journals Number: 1 Primary_atype: Research Publisher: Nature Publishing Group Subject_term: Cardiology;Machine learning Subject_term_id: cardiology;machine-learning, p. 10949. ISSN: 2045-2322. DOI: 10.1038/s41598-021-90285-5. URL: https://www.nature.com/articles/s41598-021-90285-5 (visited on 09/01/2021).

[21] Dongdong Zhang et al. “Interpretable deep learning for automatic diagnosis of 12-lead electrocardiogram”. en. In: iScience 24.4 (Apr. 2021), p. 102373. ISSN: 2589-0042. DOI: 10.1016/j.isci.2021.102373. URL: https://www.sciencedirect.com/science/article/pii/S2589004221003412 (visited on 06/04/2022).

[22] Inês Neves et al. “Interpretable heartbeat classification using local model-agnostic explanations on ECGs”. en. In: Computers in Biology and Medicine 133 (June 2021), p. 104393. ISSN: 0010-4825. DOI: 10.1016/j.compbiomed.2021.104393. URL: https://www.sciencedirect.com/science/article/pii/S0010482521001876 (visited on 06/04/2022).

[23] Ganeshkumar M. et al. “Explainable Deep Learning-Based Approach for Multilabel Classification of Electrocardiogram”. In: IEEE Transactions on Engineering Management (2021). Conference Name: IEEE Transactions on Engineering Management, pp. 1–13. ISSN: 1558-0040. DOI: 10.1109/TEM.2021.3104751.

[24] Apoorva Srivastava et al. “A deep residual inception network with channel attention modules for multi-label cardiac abnormality detection from reduced-lead ECG”. en. In: Physiological Measurement (2022). ISSN: 0967-3334. DOI: 10.1088/1361-6579/ac6f40. URL: http://iopscience.iop.org/article/10.1088/1361-6579/ac6f40 (visited on 06/04/2022).

[25] Erick A. Perez Alday et al. “Classification of 12-lead ECGs: the PhysioNet/Computing in Cardiology Challenge 2020”. en. In: Physiological Measurement 41.12 (Dec. 2020). Publisher: IOP Publishing, p. 124003. ISSN: 0967-3334. DOI: 10.1088/1361-6579/abc960. URL: https://dx.doi.org/10.1088/1361-6579/abc960 (visited on 10/26/2022).

[26] Matthew A Reyna et al. “Will Two Do? Varying Dimensions in Electrocardiography: The PhysioNet/Computing in Cardiology Challenge 2021”. In: 2021 Computing in Cardiology (CinC). Vol. 48. ISSN: 2325-887X. Sept. 2021, pp. 1–4. DOI: 10.23919/CinC53138.2021.9662687.

[27] Bjørn-Jostein Singstad and Christian Tronstad. “Convolutional Neural Network and Rule-Based Algorithms for Classifying 12-lead ECGs”. en. In: Dec. 2020, pp. 1–4. DOI: 10.22489/CinC.2020.227. (Visited on 01/22/2021).

[28] Bjørn-Jostein Singstad and Pål Haugar Brekke. “Multi-label ECG classification using Convolutional Neural Networks in a Classifier Chain”. In: Computing in Cardiology (2021). In Review.

[29] Feifei Liu et al. “An Open Access Database for Evaluating the Algorithms of Electrocardiogram Rhythm and Morphology Abnormality Detection”. en. In: Journal of Medical Imaging and Health Informatics 8.7 (Sept. 2018), pp. 1368–1373. DOI: 10.1166/jmihi.2018.2442. (Visited on 08/29/2020).

[30] Vikto Tihonenko et al. “St Petersburg INCART 12-lead arrhythmia database”. In: PhysioBank PhysioToolkit and PhysioNet (2008).

[31] Patrick Wagner et al. “PTB-XL, a Large Publicly Available Electrocardiography Dataset”. In: Scientific Data 7.1 (May 2020), p. 154. DOI: https://doi.org/10.1038/s41597-020-0495-6. (Visited on 08/31/2020).

[32] R. Bousseljot, D. Kreiseler, and A. Schnabel. “Nutzung der EKG-Signaldatenbank Cardiodat der PTB über das Internet”. In: Biomedizinische Technik/Biomedical Engineering (July 2009),pp. 317–318. DOI: 10.1515/bmte.1995.40.s1.317. (Visited on 10/18/2020).

[33] Jianwei Zheng and et al. “Optimal Multi-Stage Arrhythmia Classification Approach”. en. In: Scientific Reports 10.1 (Feb. 2020), p. 2898. DOI: 10.1038/s41598-020-59821-7. (Visited on 08/30/2021).

[34] Christian Szegedy et al. Going Deeper with Convolutions. 1409.4842 [cs]. Sept. 2014. URL: http://arxiv.org/abs/1409.4842 (visited on 09/09/2022).

[35] Martín Abadi et al. “TensorFlow: Large-Scale Machine Learning on Heterogeneous Distributed Systems”. In: 1603.04467 [cs] (Mar. 2016). arXiv: 1603.04467. URL: http://arxiv.org/abs/1603.04467 (visited on 01/31/2021).

[36] David E. Rumelhart, Geoffrey E. Hinton, and Ronald J. Williams. “Learning representations by back-propagating errors”. en. In: Nature 323.6088 (Oct. 1986). Number: 6088 Publisher: Nature Publishing Group, pp. 533–536. ISSN: 1476-4687. DOI: 10.1038/323533a0. URL: https://www.nature.com/articles/323533a0 (visited on 01/15/2021).

[37] Quintin van Lohuizen. “Training Deep Neural Networks with Soft Loss for Strong Gravitational Lens Detection”. en. MA thesis. University of Groningen. URL: https://www.ai.rug.nl/~mwiering/Thesis_Quintin_van_Lohuizen.pdf.

[38] Diederik P. Kingma and Jimmy Ba. “Adam: A Method for Stochastic Optimization”. In: 1412.6980 [cs] (Jan. 2017). arXiv: 1412.6980. URL: http://arxiv.org/abs/1412.6980 (visited on 01/30/2021).

[39] Connor Shorten and Taghi M. Khoshgoftaar. “A survey on Image Data Augmentation for Deep Learning”. In: Journal of Big Data 6.1 (July 2019), p. 60. ISSN: 2196-1115. DOI: 10.1186/s40537-019-0197-0. URL: https://doi.org/10.1186/s40537-019-0197-0 (visited on 12/05/2021).

[40] Marzyeh Ghassemi, Luke Oakden-Rayner, and Andrew L Beam. “The false hope of current approaches to explainable artificial intelligence in health care”. en. In: The Lancet Digital Health 3.11 (Nov. 2021), e745.#x2013;e750. ISSN: 2589-7500. DOI: 10.1016/S2589-7500(21)00208-9. URL: https://www.sciencedirect.com/science/article/pii/S2589750021002089 (visited on 10/28/2022).

